# Social determinants of adult COVID-19 vaccine acceptance and uptake in a Brazilian urban informal community: a longitudinal time-to-event study

**DOI:** 10.1101/2023.11.28.23298927

**Authors:** Murilo Dorión, Juan Pablo A. Ticona, Mariam O. Fofana, Margaret L. Lind, Nivison Nery, Renato Victoriano, Ananias S. do Aragão Filho, Mitermayer G. Reis, Federico Costa, Albert I. Ko

**Affiliations:** Department of Epidemiology of Microbial Diseases, Yale School of Public Health, New Haven, CT, United States; Instituto de Saúde Coletiva, Universidade Federal da Bahia, Salvador, BA, Brazil; Instituto Gonçalo Moniz, Fundação Oswaldo Cruz, Ministério da Saúde, Salvador, BA, Brazil; Faculdade de Medicina da Bahia, Universidade Federal da Bahia, Salvador, BA, Brazil

## Abstract

Residents of informal urban settlements have a high risk of COVID-19 exposure and have less access to medical care, making vaccine-driven prevention critical in this vulnerable population. Despite robust vaccination campaigns in Brazil, vaccine uptake and timing continue to be influenced by social factors and contribute to health disparities. To address this, we conducted a sequential survey in a cohort of 717 adults in an urban *favela* in Salvador, Brazil where participants were interviewed in 2020, before vaccines were rolled out, and in 2022, after primary and booster dose distribution. We collected data on demographics, social characteristics, and COVID-19 vaccination status and intent. Primary series uptake was high (91.10% for 1^st^ dose and 94.74% for 2^nd^ dose among eligible); however, booster uptake was lower (63.51% of eligible population) at the time of the second interview, suggesting a decreasing interest in vaccination. To account for both vaccine refusal and delays, we conducted a Cox time-to-event analysis of dose uptake using sequential independent outcomes. Exposure times were determined by dose eligibility date to account for age and comorbidities. Intent to vaccinate in 2020 (hazard ratio [HR]: 1.54, CI: [1.05, 1.98]) and age (HR: 1.27, CI: [1.01, 2.08]) were associated with higher vaccination rates for the 1^st^ dose. Males were less likely to receive the 1^st^ dose (HR: 0.61, CI: [0.35, 0.83]), and, compared to catholics, 2^nd^ dose uptake was lower for those identifying with Pentecostalism (HR: 0.49, CI: [0.37, 0.66]) and without a religion (HR: 0.49, CI: [0.37, 0.66]), with the latter association disappearing after controlling by age. Risk perception was associated with 2^nd^ dose uptake (HR: 1.15, CI: [1.08, 1.26]). The role of sex and religion in vaccination behavior highlights the need for targeted outreach and interfacing with local organizations. The data offers lessons to build a long-term COVID-19 vaccination strategy beyond availability.

## Introduction

As periodic COVID-19 revaccination becomes the primary tool to mitigate the effects of endemic SARS-CoV-2 spread^1,2^, addressing obstacles to timely vaccine uptake is critical. Vaccination is a complex health behavior mediated by psychological and cognitive mechanisms, socioeconomic context, and cultural factors^3–5^. When countries and local governments mitigate supply and access issues, factors as varied as medical provider opinions, perception about disease severity, and past experiences with vaccine administration can affect an individual’s willingness to receive an immunization^6^. Given the complexity of underlying mechanisms, it is unsurprising that the prevalence and social distribution of COVID-19 vaccine hesitancy varies drastically across societies^7^ and between groups within the same country^8^. This heterogeneity may create unexpected directionalities in the effect of socioeconomic factors in vaccination behavior: While evidence from the United States^9^ and the United Kingdom^8^ suggests that higher socioeconomic status (SES) increases the likelihood of vaccination, the reverse is true for ethnically Arab individuals in the national Israeli healthcare dataset, as uptake in this community decreases with lower income^10^. Thus, it is critical to understand the drivers of vaccination in a culturally and socially specific context to effectively identify and respond to coverage gaps and prioritize outreach resources.

Once Brazil rolled out the COVID-19 vaccines, the country was especially well-positioned to achieve high population coverage across social strata. Since the initial implementation of the Unified Healthcare System (*Sistema Único de Saúde,* SUS) in 1985, childhood vaccine coverage has increased across all income groups, with a longitudinal household-based survey showing that all income brackets reached 95% coverage in 1993. However, in a follow-up household-based survey conducted in 2015, the wealthiest individuals were least likely to have full vaccine coverage^11^. The consolidation of the SUS, expansion of community health programs^12^ and vaccine requirements for accessing social assistance programs^13^ have driven high coverage in lower SES groups in Brazil. On the other hand, wealthier individuals have few external incentives to vaccinate; moreover, they can access private providers and demand individual choice, and their doctors are more likely to subscribe to vaccine-skeptical beliefs^11,14^.

However, despite high coverage among the poorest strata in Brazilian society, there remain significant gaps in access and uptake within this group. For instance, studies have identified significantly lower influenza vaccination coverage among the elderly residents of two *favelas* in Rio de Janeiro compared to the citywide average^15^. Another survey in *São Luís* in the state of *Maranhão* found a reduction in vaccine confidence among evangelical Christians^16^, a religious denomination rapidly expanding in working class and poor neighborhoods^17^. Finally, a household-based survey in Salvador found that women with fewer years of education were more likely to express hesitancy about the vaccine^18^. Except for one study^15^, these results came from citywide surveys that did not specifically focus on low-income groups. Rather, uptake literature has either centered^11,14^ or recommended further academic attention^11,15,19^ on upper and middle classes, creating an important evidence gap among lower-income groups.

To address this, the present study interrogates vaccination uptake and timing in an informal urban settlement area, popularly known as a *favela*, in the São Marcos/Pau da Lima neighborhoods of Salvador, Bahia, Brazil. This community is in a peripheral area that grew informally in the 1970s after the expulsion of poor populations from the city center. The need to rely on public transit and high residential crowding have made residents especially vulnerable to COVID-19 infection^20^. However, as of 2020, intent to take the vaccine among residents was only 66.0%^21^, lower than the national estimate of 85.4%^21^ or the city-wide estimate for Salvador of 81.4%^18^. Since intent to vaccinate is not a straightforward predictor of actual COVID-19 vaccination^22^ and adult booster mandates are expected to relax, the present study addresses the pressing need to better understand vaccination behavior in this population. We used a longitudinal household-based survey and retrospective vaccination data to understand factors driving dose delays and vaccine uptake. This can inform outreach campaigns and build long-term resiliency in this vulnerable population.

## Methods

### Survey and data collection

A household-based survey was administered by trained interviewers to a closed cohort of adult residents of the São Marcos and Pau da Lima neighborhoods in Salvador, Brazil. A study area census was conducted, and all residents above 18 years were eligible to participate. Individuals who consented were interviewed twice, first between August 12, 2020 and February 26, 2021 and subsequently between March 25, 2022 and September 4^th^, 2022. The field team performed multiple house visits until residents were found or the data collection period concluded. During their first interview, participants were asked about their socio-demographic characteristics, history of chronic illness and quality of life, perceptions of COVID-19 severity, and intention to receive a vaccine if one was offered in the future. Participants responded to a similar questionnaire during their second interview and provided their COVID-19 vaccination history. All data was collected electronically using the RedCap platform^23,24^. For individuals who reported receiving at least one dose, vaccination status was verified using vaccination cards or, in the absence of physical documentation, the municipal vaccination database. Further details on the survey design and data collection can be found elsewhere^21^.

### Loss-to-follow-up adjustment

We accounted for loss-to-follow-up using the inverse probability score of treatment weighting (IPTW) resampling procedure^25^. Briefly, we used a logistic regression with a generalized linear model to predict follow-up using data from the first survey (0 for lost-to-follow-up, 1 for retained) using sex, age, intent to vaccinate, COVID risk perception, and number of afternoons spent at home. We imputed missing data using multiple imputation with the MAMI package^26^ in R, and the score was calculated from the average of the propensity scores generated by five imputed datasets. The model found that the number of afternoons at home, intent to vaccinate, age, and sex were significant predictors of a successful follow-up (Supplemental Figure S2). The scores were then used as weights to resample the original dataset with replacement, and the resulting dataset was used for modeling. After resampling, the same analysis failed to yield statistically significant coefficients, indicating the mitigation of loss-to-follow-up bias associated with these variables.

### Factor selection and preparation

Factors were considered for potential inclusion in the model from three domains. Demographic variables included age, per capita household income, religious affiliation, and sex as recorded on government identification. Race was not included as a predictor due to the overwhelming majority (94.2%) of respondents identifying as either *Preto* [Black] or *Pardo* [Brown]. In this community, both categories refer to African ancestry, and the boundaries between them are fluid, individual, and politically contested even within the study area, thus making them poor candidates to have an identifiable effect. A second set of factors was selected to assess the pathway to vaccination and the attitudes and environmental factors that affect access to vaccination (Figure 1B). Intention to get vaccinated (assessed in 2020 prior to availability of vaccines) and COVID-19 risk perception measured in 2022 are related to the intent (motivation) to get vaccinated, while afternoon spent at home and employment status serve as proxies for free time (environmental constraints). Health status was assessed using the SF12 score and likely affects motivation and constrain action. Participation in the *Bolsa Família/Auxílio Brasil* cash transfer program was included in the model due to the previously documented effects of the program on health behavior and vaccination^27,28^. The model also included effect modification between intent and sex, afternoons and sex, age and physical health, age and religious identity, and mental and physical health.

Vaccine eligibility dates were estimated using an individual’s age and recorded comorbidities, which were cross-referenced with the municipal health department’s vaccine rollout announcements. Since local authorities sometimes started priority group vaccination a few days early, individuals who got vaccinated up to 14 days before their official eligibility had their eligibility date brought forward to coincide with their actual vaccination date. The SF12 scores for physical and mental health were calculated from survey responses using the procedure described in Ottoboni et al.^29^. All numeric variables were scaled to have mean 0 and standard deviation of 1 for the modeling analysis. Participants were considered as participating in the *Bolsa Família/Auxílio Brasil* cash transfer program if at least one person in the household reported being a beneficiary, as the program is family-based rather than individual. For religious affiliation, Afro-Brazilian religions, Spiritism and Other were collapsed into a single “Other” category because, although they are culturally distinct and have a significant presence in the community, the low number of participants reporting affiliation to each, and the large number of religious categories in the data, could cause model saturation. Missing data was handled by multiple imputation on the resampled dataset (K=5)^30^.

### Statistical approach

The 3-dose vaccination series was conceptualized as a sequential process, fitting the completion of each dose with a Cox Proportional Hazards model (A 4^th^ dose started being rolled out after the start of data collection, so it was excluded from the analysis). For any dose in the series, the outcome of each model was defined as the date of vaccination with a given dose, and a participant was censored at the date of the interview if their vaccination records did not show a dose at that time. Since individuals are not at risk of receiving a given dose before they become eligible or before receiving the prior dose, the model was defined as a 6-state model, and the relevant coefficients correspond to the transition between dose eligibility and completion of the dose (see figure 1A).

Coefficient estimation for each model in the vaccination sequence can be performed independently when the timing of receiving a dose is not correlated with delays in previous doses (Markov assumption); however, such a premise can be too restrictive for vaccination phenomena. For this reason, instead of using calendar time, the models were fit using a semi-Markov timescale in which time equals zero whenever someone starts being at risk for a new outcome^31^. The Markov assumption was tested after the adjustment using the procedure recommended in Cook and Lawless^31^ and, if the process still did not conform to the Markov assumption, we included the calendar time of entry into eligibility as a predictor as described by Andersen, Wandall, and Pohar^32^ to model non-Markov Cox processes.

The model was fit, and the coefficients were chosen, with AIC model selection using the MAMI package. The five imputed datasets were each used to select a model using both forward and backward stepwise selection, and the point estimates were derived by averaging the five models^26^. In this framework, a variable is selected if it appears in the model from at least one of the multiply imputed datasets, but the effect size depends on the number of times it is selected. To accurately estimate imputation and variable selection uncertainty, a bootstrapping procedure was used where the multiple imputation (K=5) and the model selection algorithm were repeated for 200 subsamples of the main dataset. Coefficients are reported in the study if they appeared in at least 95% of bootstrap model selections, and the confidence intervals were derived from the empirical bootstrap distribution of the coefficient estimates from all models that selected the variable. A comparison between point estimates from the base model (used in the analysis) and bootstrapping averages is included in the appendix.

When the reported model result violated the proportional hazards (PH) assumption in the complete case, categorical time-dependence was allowed for the violating variable by creating a binary time variable with a cutoff time chosen using the model residuals (100 days for 1^st^ dose, 30 days for 2^nd^ dose and 40 days for 3^rd^ dose). For age in 2nd dose model and physical health in 3rd dose, the PH violation persisted despite the inclusion of these factors in time-varying manners, and the variable was thus dropped to preserve model integrity. In both cases, the model including the problematic variable is included in the supplement (Supplemental Figure S3).

## Results

Of the 1956 eligible residents, 1130 (57.7%) were initially included in the cohort, and 717 adults responded to both interviews (63.4% follow-up rate; see Table S1 for demographics). Among the 1130 adult residents who initially consented, 949 (63.6%) stated that they intended to take a COVID-19 vaccine if one was offered, while 400 (26.8%) stated that they did not intend to take the vaccine, and 142 (9.5%) were not sure. A detailed description of the initial survey results can be found elsewhere^21^. Among the 717 who responded to both interviews, 491 (69.7%) stated that they intended to take the vaccine, while 162 (22.5%) did not intend and 50 (7.0%) were not sure. The bias from the high loss-to-follow-up rate (36.6%) was addressed using IPTW resampling (see Methods).

Vaccination coverage was high for the primary series, with 91.90% of eligible individuals reporting 1st dose uptake and 93.81% with a verified 2^nd^ dose. However, 3^rd^ dose uptake was lower, with only 63.51% of the eligible population having taken the first booster dose at the time of the interview (Figure 2).

**Figure 1:**
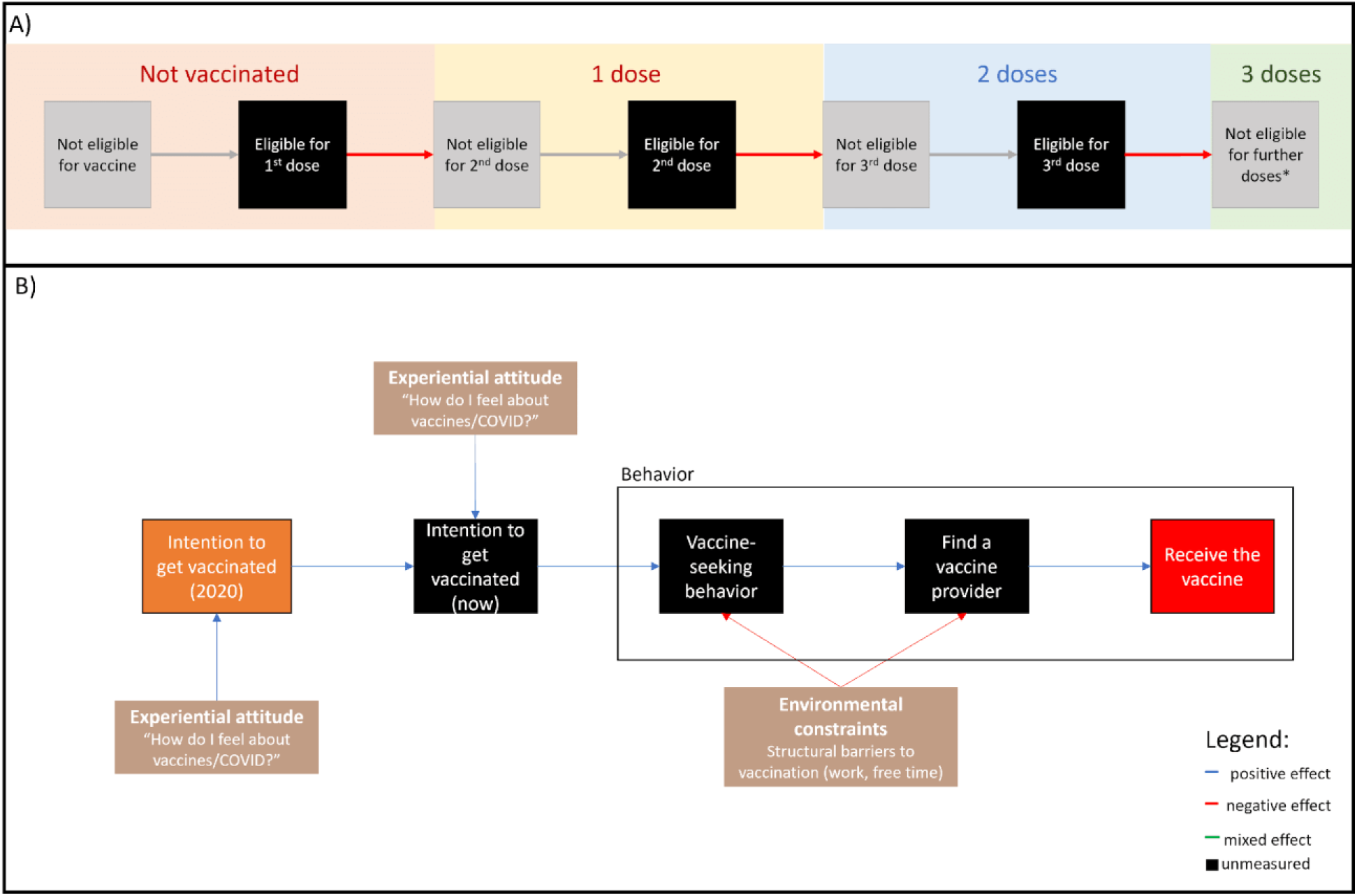
schematic representation of modeling approach. *(A) Representation of vaccination pathway as a sequential process.* The diagram shows the ascertained status (dose number, background) and the hidden state (squares showing eligibility status for the following dose). The time in which an individual is eligible (solid black), and thus at risk of a vaccination event (red arrows), is the only state that contributes person-time to the model. Separate independent models are developed for each vaccination event as described in the text. (B) Conceptual model used for *a priori* feature selection. This model is an adaptation of the Integrated Behavioral Model (IBM)^3^ applied to vaccination. The core pathway shows the chain of events that ultimately leads to receiving the vaccine. First, intent to vaccinate in 2020 affects the present (unmeasured) intent to vaccinate. Motivation then propels individuals to take action in the form of vaccine seeking, which can result in successfully finding a vaccine provider and, if the engagement is successful, they receive a vaccine. Intent to vaccinate is affected by perceptions of disease severity, and the pathway between intent to vaccinate and receiving the dose is mediated by environmental constraints.

**Figure 2:**
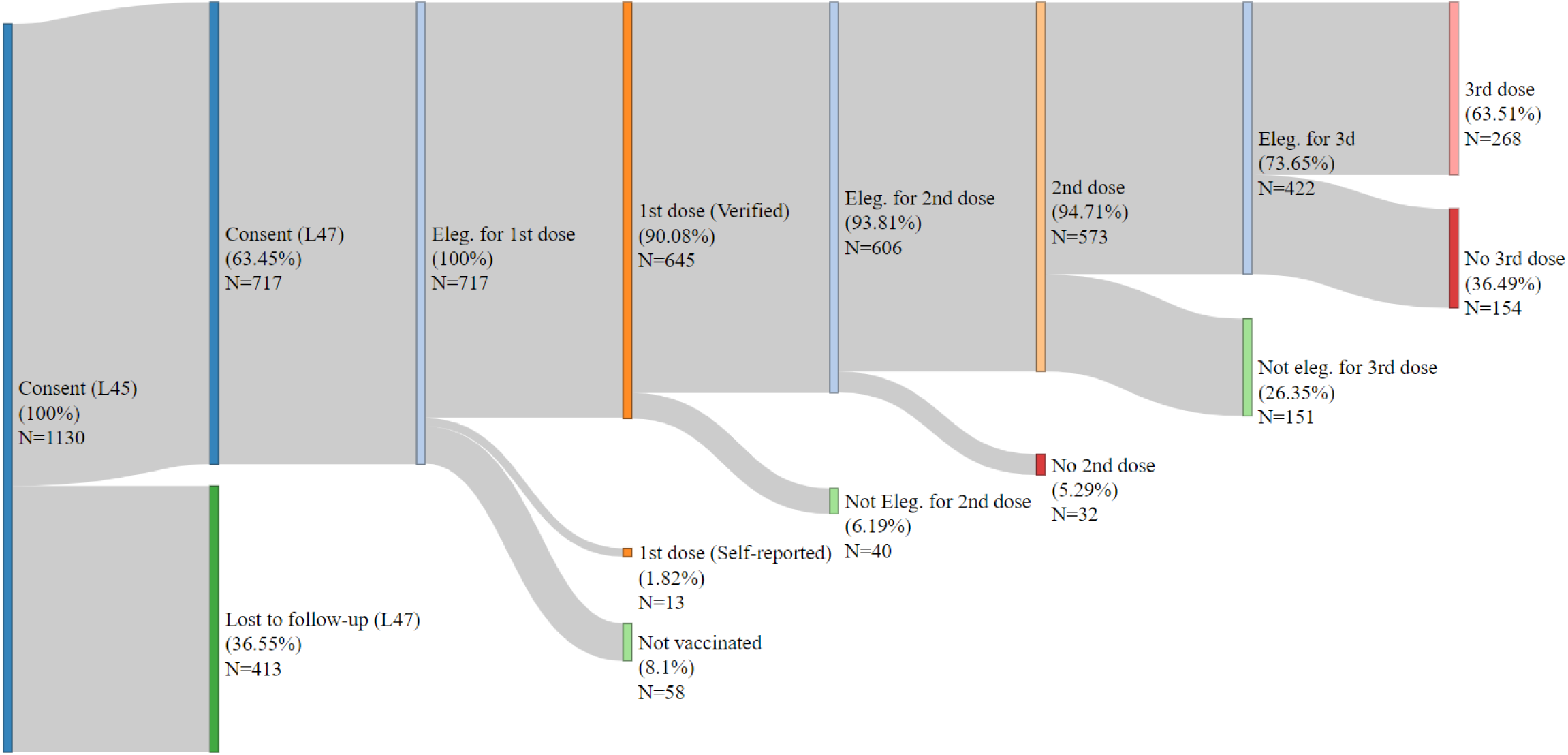
Sankey diagram of adult vaccination status and eligibility at the time of interview. Percentages are calculated with the previous stage as the denominator, so vaccination percentages refer to the eligible population. Self-reported vaccination histories that could not be verified in the municipality system (N=11) were excluded from the analysis due to the unreliability of precise vaccination date recall.

### Vaccination rollout

Retrospective vaccination data shows an initial vaccination uptake followed by a plateau in primary series uptake (Figure 3A). For the 1^st^ dose, we observed a rapid increase in vaccine uptake at the start of vaccination, but the rate slowed by mid-August/21. The 2^nd^ dose curve tracks 1^st^ dose uptake, with faster uptake between August/21 and December/21. However, The third dose coverage did not plateau, having increased approximately constantly since the initial rollout in November/22.

**Figure 3:**
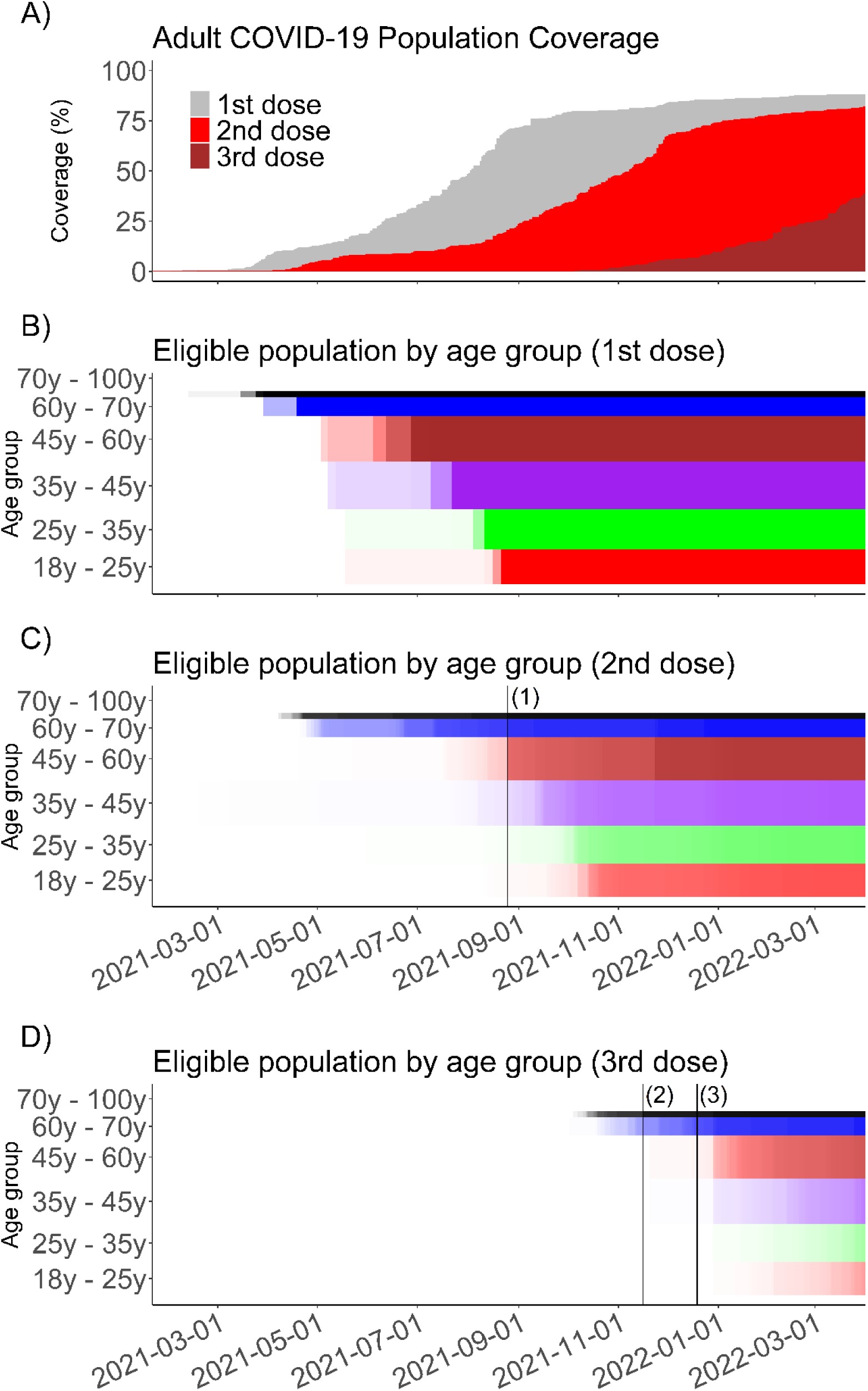
Summary of vaccination rollout in the community. *(A) Total vaccination coverage among participants by dose between January 18, 2021 and April 1^st^, 2022.* The data shows a gradual increase in 1^st^ dose coverage followed by a plateau starting around November 2021. For 2^nd^ dose coverage, there is an initial, slower total uptake before late July, followed by a steeper total coverage increase until December 2021. Third dose coverage is constantly increasing between November and the interview date, and it had not yet reached a plateau. (B) Visualization of total 1^st^ dose eligibility by age group. Each horizontal bar represents an age group, labeled on the Y-axis, and the opacity of the bar is proportional to the percentage of each age group that is eligible for the 1^st^ dose at a given time. The height of each bar is proportional to the share of the study population that belongs to a given age band, so a larger band represents a larger group. (C) Visualization of 2^nd^ dose eligibility by age group. Second dose eligibility trails 1^st^ dose uptake on all age groups. Vertical line (1) marks the date when the interval between 1^st^ and 2^nd^ dose eligibility for Pfizer and AstraZeneca was shortened from 12 to 8 weeks. (D) Visualization of 2^nd^ dose eligibility by age group. Vertical line (2) marks the date when the interval for 3^rd^ dose eligibility was reduced from 180 to 150 days, while line (3) represents the further reduction from 150 days to 120 days.

**Figure 4:**
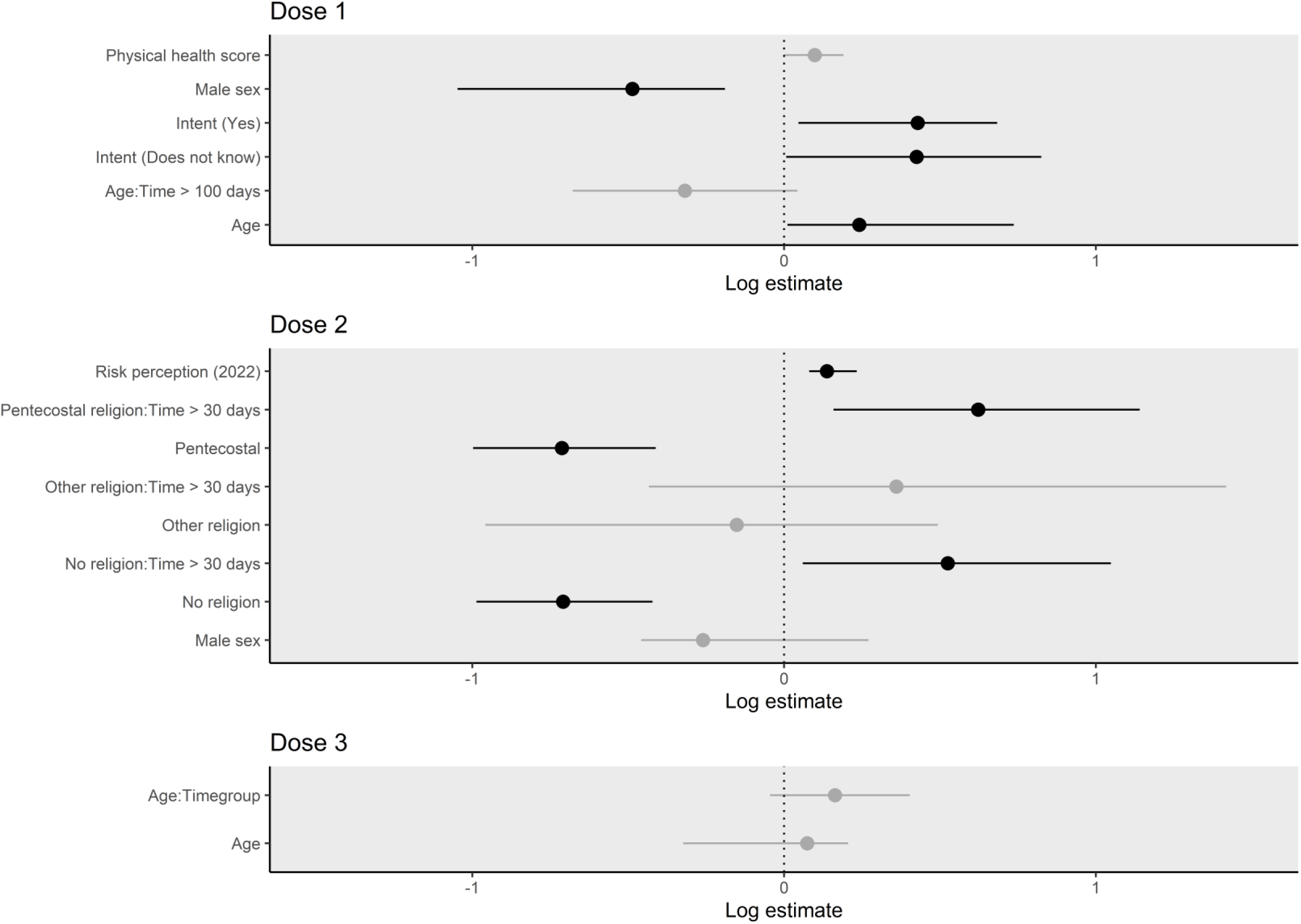
Model results. Log coefficient estimates are included if the variable was selected in at least 95% of bootstrapped models, and error bars represent the 95% bootstrap interval (B=200).

Some participants between 25 and 60 years became eligible before age-based eligibility due to their profession or comorbidities, but most became eligible together with their age group (Figure 3B). The 1^st^ dose uptake curve also tracks the increase in the eligible population, with the uptake plateau coinciding with the timeframe when all adults became eligible to vaccinate. A similar behavior is observed for the 2^nd^ dose (Figure 3C), with faster uptake occurring after a policy change which reduced the interval between the first and second dose. Eligibility for 3^rd^ dose is concentrated in 2022 for most age groups, despite previous shortenings of waiting intervals (Figure 3D).

### Factors associated with vaccine uptake

Figure 4 shows the sequential Cox regression coefficients. For the 1^st^ dose, compared to those who did not intend to get vaccinated (ref.), the hazard of vaccination was 1.54 (95% Confidence Interval [CI]: 1.05, 1.98) times higher for people who were intending to vaccinate in 2020, and 1.78 (CI: 1.29, 2.87) for those who were not sure about taking the vaccine at that time. Relative to female participants, males were less likely to be vaccinated (HR: 0.61, CI: [0.35, 0.83]). Age was also found to be significantly associated with 1^st^ dose uptake (HR: 1.27, CI: [1.01, 2.08]).

For the 2^nd^ dose, Pentecostals (HR: 0.49, CI: [0.37, 0.66]) and those without a religion (0.49, CI: [0.37, 0.66]) had a lower risk of vaccination within 30 days of eligibility when compared to Catholics (ref.). After 30 days, however, both Pentecostals (HR: 1.86, CI: [1.17, 3.13]) and those without a religion (HR: 1.69, CI: [1.06, 2.85]) had an increased vaccination modifier when compared to others. For the sensitivity analysis including age in the model, the time-invariant effect of no religion and both time-varying coefficients ceased to be statistically significant. Pentecostal retained a significant time-invariant negative coefficient. Risk perception was found to be independently correlated with 2^nd^ dose uptake (HR: 1.15, CI: [1.08, 1.26]). No statistically significant associations were observed for the 3^rd^ dose.

## Discussion

Despite low intent to vaccinate in this cohort, which was as low as 66% in 2020^21^, primary series coverage was high in the study population (91.9% for 1^st^ dose and 94.74% for those eligible for the 2^nd^ dose). However, there is a decreasing uptake signal for the 3^rd^ dose, with only 63.51% of those eligible taking the dose. It is also concerning that, while uptake closely follows eligibility for the 1^st^ and 2^nd^ dose (Figure 3), the early-adoption behavior did not repeat for the 3^rd^ dose. This may be explained by the fact that, at the time of interview, access to governmental and some religious buildings required proof of primary series vaccination but not booster uptake, thus reducing the incentives for timely uptake.

Intent to vaccinate in 2020 is associated with 1^st^ dose uptake (HR: 1.54, CI: [1.05, 1.98]), but not for other doses. The lack of an association between intent to vaccinate and 2^nd^ and 3^rd^ dose uptake may indicate that incentives to take the vaccine and perceptions about immunization evolved throughout the pandemic, corroborating previous studies that show that vaccine hesitancy does not perfectly predict uptake^22^. These results highlight the importance of longitudinal vaccination studies to understand immunization behavior: Intent to vaccinate after vaccination can be affected by a post hoc rationalization of the vaccination decision^4^ but, by measuring intent before the rollout, this study was able to show the decreasing importance of pre-rollout intent on actual behavior. The data also suggests that the communication and incentive structure in Salvador, with information campaigns, door-to-door outreach, and limited mandates, successfully drove behavior during the pandemic phase of the disease, decreasing the importance of individual intent for timely uptake.

However, as incentive measures are relaxed, an essential component of long-term coverage resiliency is disease risk perception, which was significantly associated with 2^nd^ dose uptake (HR: 1.15, CI: [1.08, 1.26]). This presents an avenue for action as well as a challenge for maintaining high coverage. A higher initial vaccine coverage can decrease population-level disease severity, thus reducing individual’s risk perception and lowering the drive to vaccinate^2^. However, the perception that a health behavior will yield benefit is also a critical driver for action^3^. Thus individuals must believe that the disease protection is worth the cost of seeking a vaccine and the potential side effects. Effectively communicating the benefits of vaccination while emphasizing the risks posed by the disease will continue to be an integral component of an endemic-phase vaccination campaign.

People with Male sex in their official document were found to have lower hazard of taking the 1^st^ dose (HR: 0.61, CI: [0.35, 0.83])). This is surprising in light of the prior data showing that, throughout the city, men had higher vaccine confidence than women, and non-White women with fewer years of education were more likely to express vaccine hesitancy^18^. Such a result may be related to the high level of COVID-19 exposure experienced by women in the community^20^, which could increase their propensity to seek vaccination. However, local providers also express that men exhibit less healthcare-seeking behavior beyond just vaccination. As the effect of sex is likely due to its role as a proxy for gender, further research is necessary to elucidate the gendered dynamics in informal urban communities with respect to health and gender, as specific cultural, religious, and socioeconomic practices and dynamics of discrimination can affect health behavior. Data collection on a qualitative study on the matter is ongoing.

Finally, the model shows that those who identify as Pentecostal were 0.546 (CI: [0.37, 0.75]) times, and those without a religion were 0.48, CI: [0.35, 0.69]) times as likely to take the 2^nd^ dose as Catholics (ref.) within the first 30 days. The lower vaccination odds for those who do not identify with a religion may be associated with lower social capital, as religious groups are vital socialization venues in the community; however, this result is partly explained by the younger age of those in this group, as the effect ceases to be significant when accounting for age (Supplemental Figure S3). However, the lower vaccination odds among Pentecostals persists even when including age in the model. Evangelical Pentecostals, who were the vast majority (99.1%) of Pentecostals in our cohort, were also found to have lower vaccine confidence in another major Northeastern city^16^, underscoring the need for an improved understanding of the impact of this growing religion on vaccination. Brazil^33^ and Latin America in general^34^ have been undergoing a religious transition away from Catholicism, with Pentecostalism increasing in numbers and cultural importance. The impact of religion on vaccination behavior is complex, as it encompasses beliefs, moral values, tightly knit social networks, and past experiences and identities that lead individuals to follow a particular religion. A better understanding of religion’s impact on vaccination in vulnerable communities is critical to better design outreach campaigns without stigmatizing those cultural groups, and to elucidate effective ways to interface with faith-based organizations and drive behavior in these communities.

The results of the study, however, are not without limitations. Although household-based surveys are better than electronic health record data at providing a reasonable estimate of the total population when estimating vaccination coverage, they may bias the data toward more easily located respondents. Throughout the multiple months of data collection for each survey, the field team has attempted to mitigate participant selection biases by revisiting homes until they interview every resident, scheduling house visits ahead of time, and having a field schedule that included the weekends. However, some bias may remain. Despite using IPTW to mitigate loss-to-follow-up issues, the high loss rate may exacerbate this convenience bias. The study also failed to detect an effect from the “Other” religious category. As explained in Methods, this level collapsed various diverse religious traditions, so the lack of an identifiable effect is expected. Although this study was underpowered to address the relationship between Spiritism and Afro-Brazilian religions and vaccination, the results from this study highlight the importance of religion in vaccination behavior, so further research on these faiths is critical. Further, inability to detect a 3^rd^ dose effect may have been caused by the relatively short 3^rd^ dose follow-up time a large part of the study cohort, as the rollout was still ongoing during data collection. Therefore the model did not have enough person-time to adequately power the coefficient estimates. Updated analyses with longer follow-up time are ongoing.

This study provides timely evidence to inform outreach and adult vaccination campaigns in informal urban settlements. The high coverage for primary series and the lack of an association between intent to vaccinate in 2020 and 2^nd^ and 3^rd^ dose uptake are both encouraging results and underscore the importance of well-designed incentives and outreach. However, improvements are needed to interface with gender and religion regarding vaccines. Further research is crucial to better understand vaccine behavior in urban informal settlements. However, in the meantime, interfacing with community organizations is an essential tool to reach those groups locally and create resilient vaccine uptake systems.

## Ethical considerations

The Institutional Review Board of the Instituto Goncalo Moniz, Oswaldo Cruz Foundation (Fiocruz), the Brazilian National Commission for Ethics in Research (CAAE 35405320.0.1001.5030), and the Yale University Human Research Protection Program (2000031554) gave ethical approval for this work.

## Data Availability

Limited data to reproduce key results is available upon reasonable requests to the authors.

## Acknowledgements

We are deeply thankful to the members of the field team, listed alphabetically: Adrielle Nascimento, Aline Miranda, Bianca Marques, Crislane Lopes, Daniela Higino, Elaine Ferreira, Emily de Souza, Jackson dos Santos, Jaiane de Santana, Janaina Lima, Jessica Clementino, Juliana Luz, Laiara dos Santos, Leila Santos, Luana Santana, Lucicleide de Oliveira, Milena Silva, Maria Eduarda dos Santos, Monique dos Santos, Patricia Souza, Priscila Santos, Rafaela Ribeiro, Rebeca de Mattos, Roseane Rebouças Moura, Ruan Oliveira, Simone Barbosa, Singrid Santos, Stephanie Santiago, Taiane da Silva, Tassia Santos, Vitor Pereira, and Vitoria Miranda. They worked tirelessly to locate residents, recruit participants, collect data, and curate the relationship with the community that make this work possible.

## Supplementary material

**Table S1:** participant demographics.

|  |  |
| --- | --- |
| Age (mean) | 40.63 years |
| Per capita monthly income (mean) | R\$501.63 |
| Female | 63.3% |
| Religion: Catholic | 24.25% |
| Religion: Pentecostal | 31.20% |
| Religion: None | 39.41% |
| Religion: Afro-Brazilian | 3.69% |
| Religion: Spiritism | 0.28% |
| Religion: Other | 1.41% |
| Race: *Preto* [Black] | 54.18% |
| Race: *Pardo* [Brown] | 40.00% |
| Race: *Branco* [White] | 5.11% |
| Race: Other | <1% |

**Figure S2:**
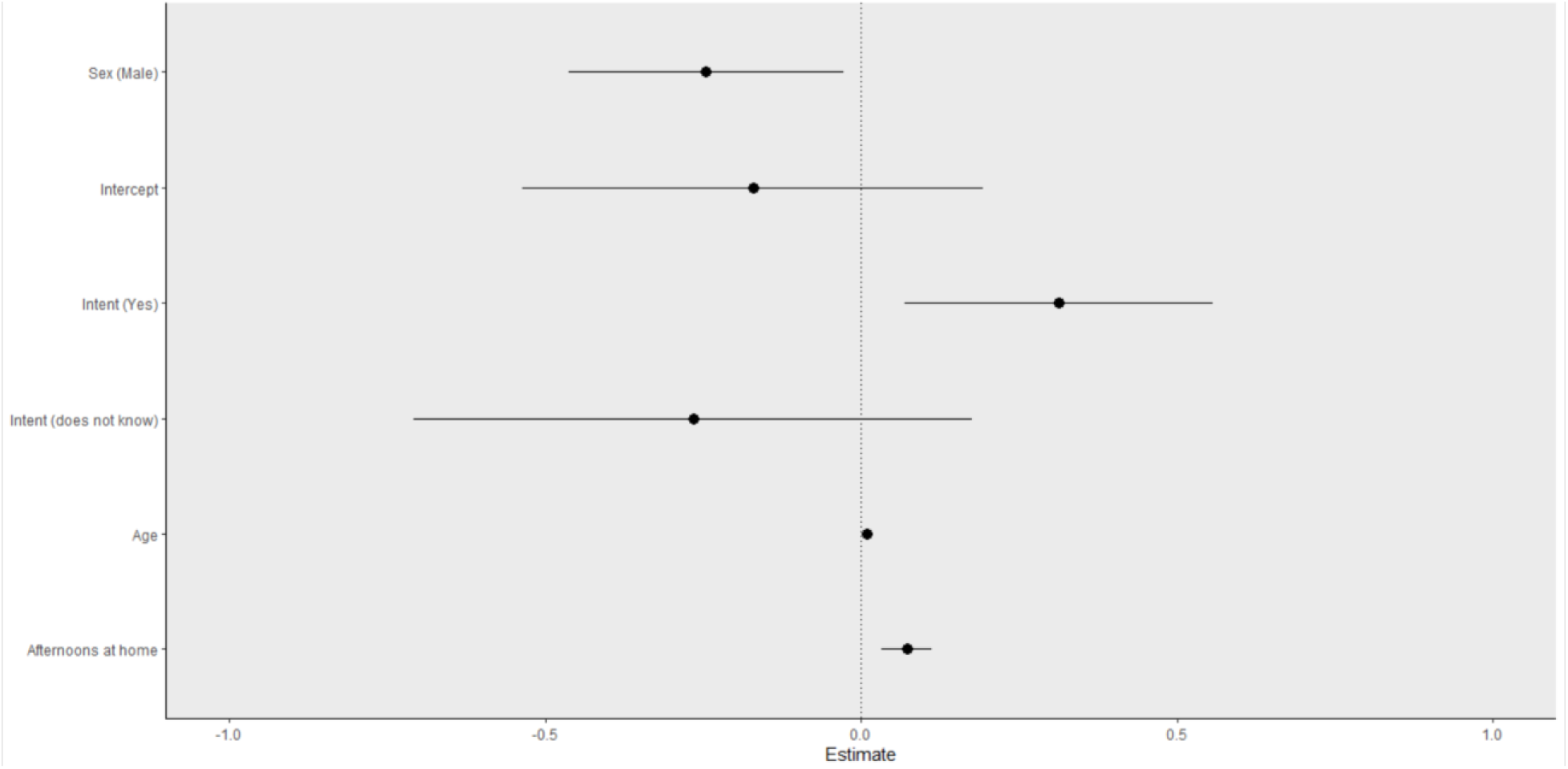
IPTW model results before resampling

**Figure S3:**
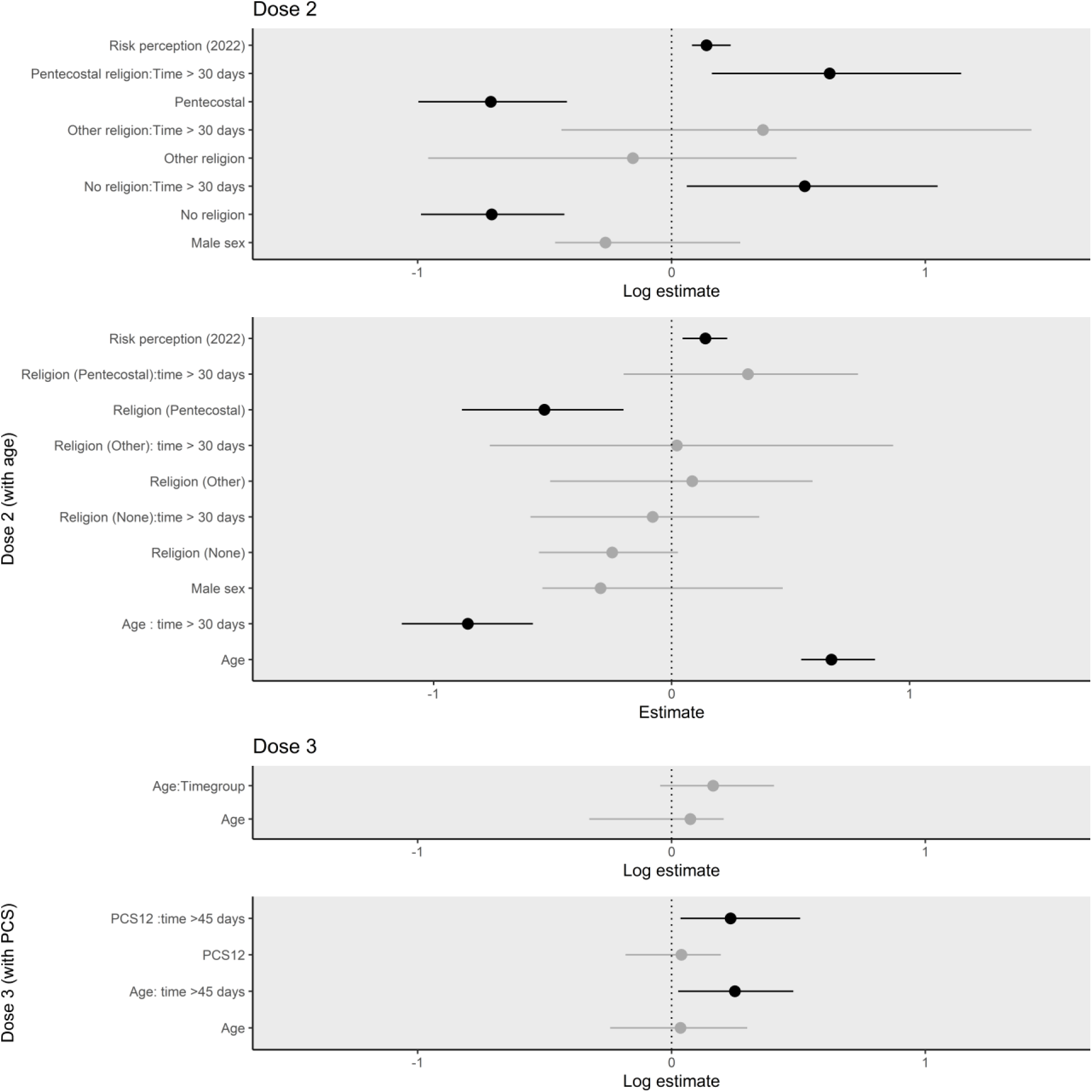
comparison between 2nd dose model with and without age, and 3^rd^ model with and without PCS12 score.

**Table S2:** Comparison between log values of the base estimate and bootstrapping average.

| Dose 1 | Variable | Estimate | Lower CI | Upper CI | (Boot mean) |
| --- | --- | --- | --- | --- | --- |
|  | Intent (Yes) | 0,43 | 0,05 | 0,68 | 0,39 |
|  | Age | 0,24 | 0,01 | 0,74 | 0,34 |
|  | Intent (Does not know) | 0,43 | 0,01 | 0,83 | 0,42 |
|  | PCS12 | 0,10 | 0,00 | 0,19 | 0,09 |
|  | Age: time > 100 days | -0,32 | -0,68 | 0,04 | -0,32 |
|  | Male sex | -0,49 | -1,05 | -0,19 | -0,54 |
| Dose 2 | Male sex | -0,26 | -0,46 | 0,27 | -0,22 |
|  | Religion (None) | -0,71 | -0,99 | -0,42 | -0,69 |
|  | Religion (Other) | -0,15 | -0,96 | 0,49 | -0,14 |
|  | Religion (Pentecostal) | -0,71 | -1,00 | -0,41 | -0,71 |
|  | Risk perception (2022) | 0,14 | 0,08 | 0,23 | 0,15 |
|  | Religion (None): time > 30 days | 0,53 | 0,06 | 1,05 | 0,50 |
|  | Religion (Other): time > 30 days | 0,36 | -0,43 | 1,42 | 0,40 |
|  | Religion (Pentecostal): time > 30 days | 0,62 | 0,16 | 1,14 | 0,61 |
| Dose 3 | Age | 0,07 | -0,32 | 0,21 | -0,01 |
|  | Age: time > 45 days | 0,16 | -0,05 | 0,40 | 0,18 |

